# Too many yet too few caesarean section deliveries in Bangladesh: an ongoing public health challenge to improve maternal and child health

**DOI:** 10.1101/2021.08.28.21262769

**Authors:** Md Nuruzzaman Khan, Md Awal Kabir, Asma Ahmad Shariff, Md Mostafizur Rahman

## Abstract

**Background:** Caesarean section (CS) use is on the rise in Bangladesh, particularly among women in improved socio-economic condition. However, the deficit use of CS remains common among disadvantage women in terms of employment, education, wealth quintile, and place of residence. This increases risks of long-term obstetric complications as well as maternal and child deaths. We aimed to determine the interaction effects of women’s disadvantage characteristics on CS use in Bangladesh.

**Methods:** Total of 27,093 women data analysed extracted from five rounds of Bangladesh Demographic and Health Survey, conducted between 2004 and 2017/18. The inclusion criteria used to select these women were: (i) having at least one child within three prior to the survey, (ii) reported delivery methods and place, and (iii) do not have twin or more ordered pregnancy for the most recent live birth. The major exposure variables were type of health facilities, divisions, place of residence, economic status, and maternal education. Other factors considered were factors at the individual and household level. The outcome variable was CS use, coded as use (1) and non-use (0). Multilevel logistic regression model was used to determine association of CS with socio-demographic characteristics and the interactions of the working status and wealth quintile with place of residence.

**Results:** We reported a 751% increase of CS use over the last 13 years — from 3.88% in 2004 to 33% in 2017/18. Nearly, 80% of these occurred in the private health facilities followed by the government health facilities (15%). Rural women with no engagement of formal income generating activity showed 11% (OR, 0.89, 95% CI, 0.71-0.99) lower use of CS in 2004. This association was further strengthened with the year passes, and a 51% (OR, 0.49, 0.03-0.65) lower in CS use was reported in 2017/18. Similarly, around 12%-83% lower likelihoods of CS use were found among rural poor and poorer women.

**Conclusion:** Bangladesh is facing a double burden of CS, that is a group of women with improved socio-economic condition using this life saving procedure without medical necessity while their counterpart of disadvantage characteristics could not access this service. Improved monitoring from the government along with support to use CS services for the disadvantage groups on necessity are important.

## Introduction

The number of caesarean section (CS) use continues to rise globally, now accounting 21% of all childbirths with a significant variation across the countries [1]. The proportion is less than 5% in 28 countries worldwide, over three quarter of them are located in sub-Saharan Africa, including Niger, Chad, Ethiopia, and Timor Laste [2]. Only 10% of the total countries worldwide have the CS rate 10-15%[3], which is the World Health Organization recommendation to the significant reductions of maternal and child mortality [4]. Over 100 countries worldwide have above 15% CS use; 43 countries even have their CS use level higher than 30%. This later group is geographically heterogeneous and mostly developed countries [2]. However, recent rise on CS rates is mainly occurring in low- and lower-middle income countries (LMICs) where improving maternal health is an ongoing challenge.

Around 42% of the total CS performed worldwide are without medical necessities, therefore they do not have any contributions in improving maternal and child health [2]. Moreover, such abusive use of CS can lead several public health burdens, including hemorrhage and bleeding as well as associated maternal mortality and economic burden [5-7]. This is also found to be linked with a long-term loss of women’s productivity as well as increasing hospitalization which further create a burden in formal healthcare delivery system [6-8].

The CS use in LMICs is generally linked to the level of development including women’s education, fertility level, wealth, and the weight of the private healthcare facilities in providing CS [9, 10]. Consequently, a group of women could not access this service because of financial hardship whereas another group use this service without medical necessity — an indication of the double burden of CS [10]. Moreover, along with financial capacity, geographical variation of availability of this service as well as geographical hardship to access services, such as poor transportation, could also play a significant role in differencing the CS use [11, 12]. This indicates triple burden of CS — a common scenario for LMICs including Bangladesh. The underlying reasons are unavailability of formal health insurance coverage, higher rate of poverty, and rurality — the issues which are quite significant in Bangladesh [2]. This leads a portion of women in LMICs, and Bangladesh could not access this service even though a higher prevalence of anemia and nutritional burden are common in these groups, therefore, they are more likely face pregnancy complications with a higher need of CS use [2, 13-16]. Existing research in LMICs and Bangladesh reported lower use of CS among poor and a few geographical regions [9, 10, 12, 17, 18]. However, such burden is mostly been ignored in LMICs with rapid rising of CS use is always in the discussion of the health researchers and policy makers [13].

Researchers in LMICs including Bangladesh have been explored several factors associated with the CS use and the raising trend of CS use [5, 9, 10, 18, 19]. A significant variations of CS use across factors, such as wealth quintile, education, working status, and urban/rural and regions have also been identified [10]. However, their interaction effects on CS use are not yet explored. Consequently, the disadvantages groups of women who could not access this life saving services are mostly unknown. This increases the associated maternal and child mortality which is a challenge in achieving Sustainable Development Goals’ target 3.7 (ensure universal coverage of sexual and reproductive health) by 2030 [20]. We conducted this study to fill this gap by determining the interaction effects of women’s disadvantage characteristics including place of residence, education, wealth quintile, working status on CS use.

## Methods

### Study setting and design

We analysed five rounds of Demographic and Health Survey (BDHS) Data, collected in 2004, 2007, 2011, 2014 and 2017/18. The National Institute of Population Research and Training conducted these surveys as part of the Demography and Health Survey Program of the USA. The questionnaire used for data collection was similar in all the surveys and the sampling strategy was unique, therefore they are comparable. The broad description of the survey sampling procedure is available elsewhere [21-25]. Briefly, these surveys were conducted using the multistage random sampling procedure. At the first stage of sampling, a fixed number of Primary Sampling Units (PSU) was selected randomly. The sampling frame prepared by the Bangladesh Bureau of Statistics as part of the National Population Census 2000 (for 2004, 2007 & 2011 surveys) and 2010 (for 2014 & 2017 surveys) were used to select the PSUs. Household listing operation in the selected PSUs was then carried out at the second stage of sampling. Finally, a fixed number of 30 households were selected to be included in the survey. The inclusion criteria were unique for all surveys: (i) ever married women aged 15-49 years who are usual residence of the selected households or (i) ever married women aged 15-49 years who passed the most recent night before the survey date in the selected households but not the usual residence. Selected women’s reproductive data such as the number of births and the use of maternal healthcare services in the most recent pregnancy were collected. Their husbands’ and under-fived aged children’s data were also included.

### Data

Total of 27,093 women data, as presented in Table 1, were extracted from these five rounds of BDHS for analysis. Women were included based on the following criteria: (i) having at least one child within three prior to the survey, (ii) reported delivery methods and place, and (iii) do not have twin or more ordered pregnancy for the most recent live birth.

**Table 1:**
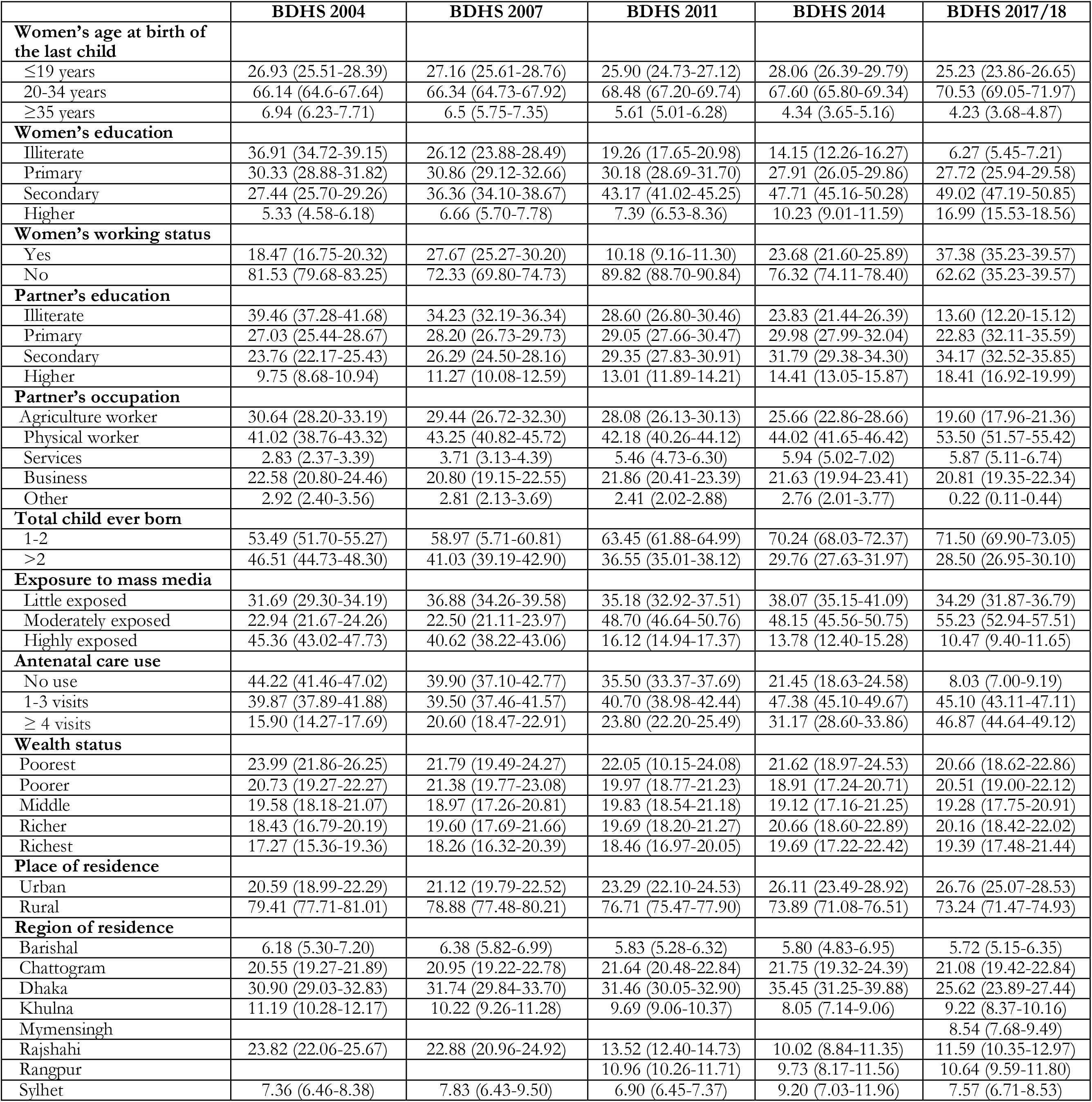
Background characteristics of the respondents, BDHS 2004-BDHS 2017/18

### Outcome variable

The outcome variable was CS use. The question asked by BDHS to the respondents: “*Was (NAME of the last child occurred in three years) delivered by caesarean, that is, did they cut your belly open to take the baby?*” The response recorded was ‘use’ (coded as 1) and ‘non-use’ (coded as 0).

### Exposure variables

The major exposure variables were type of health facilities, divisions, place of residence, economic status, and maternal education. Other factors considered were maternal age, occupation, number of children ever born, number of antenatal care visits, preceding birth interval and exposure to mass media.

### Statistical analysis

Descriptive statistics on sampling characteristics and CS use were computed for all surveys. The use of CS across type of health facilities and women’s socio-demographic characteristics were also explored. Multilevel logistic regression models were used for the association of caesarean delivery with socio-demographic characteristics at five-time points (BDHS surveys: 2004, 2007, 2011 and 2014). The reason for using the multilevel modelling was hierarchical data structure [26]. Possible pairs of interaction effects were considered in each model. If an interaction effect was found to be insignificant, the model was deleted and a new model with a different pair was run. Finally, the interactions of the working status and wealth quintile with place of residence produced significant results, therefore reported in the table. Results are reported as Odds Ratio (OR) and its 95% Confidence Interval (95% CI). The Statistical Package R were used for all statistical analyses.

## Results

The characteristics of the respondents’ analysed are presented in Table 1, separately for each survey. More than two-third of the total women analysed were in the range ages of 20-34 years. Around 37% of the women analysed in 2004 survey were illiterate which declined to nearly 6% in 2017 survey. On contrary, the percentage of higher education increased to nearly 17% in 2017 from only 5% in 2004. Over two-third of the total women did not have any formal working status in all the surveys. The total number of children ever born reported was found to be declined from 2004 to 2017. In the 2004 survey, around 53% of the total women had 1-2 children which was found to be increased to 72% in the 2017 survey. Percentage of highly exposed to mass media was found to be declined, from 45% in the 2004 survey to the 10% in 2017. ANC visit of at least 4 times was found to be increased, from only 16% in the 2004 survey to nearly 47% in the 2017 survey.

The percentage of institutional delivery, CS use, and their changes over the survey years are presented in Table 2. Around half of the total women given their last birth in healthcare institutions in 2017, increased from only 11% in 2004 — a 357% increase in 13 years. At the same time, CS use increased nearly 751%, from 3.88% in 2004 to 33.22% in 2017/18. This increase was nearly 844% for CS use among institutional delivery only, 36.83% in 2004 to 66.66% in 2017/18.

**Table 2:** Institutional delivery and caesarean delivery rates and changes in Bangladesh, 2004 to 2017/18

| Survey | Period | Number of births | Delivery rate, % (95% CI) |  |  | Percentage change across the years |  |  |
| --- | --- | --- | --- | --- | --- | --- | --- | --- |
|  |  |  | Institutional | Population caesarean | Institutional caesarean | Institutional | Population caesarean | Institutional caesarean |
| BDHS 2004 | 2001-2003 | 5361 | 10.89 (9.63-12.30) | 3.88 (3.25-4.62) | 36.13 (31.66-40.85) | -- | -- | -- |
| BDHS 2007 | 2004-2006 | 4870 | 16.36 (14.65-18.24) | 8.51 (7.41-9.76) | 52.54 (47.99-57.06) | 50.22 (48.29-53.98) | 119.33 (111.26-128.00) | 454.19 (396.82-515.79) |
| BDHS 2011 | 2006-2010 | 7272 | 26.51 (24.87-28.22) | 14.90 (13.74-16.14) | 56.89 (54.09-59.65) | 143.43 (129.43-163.33) | 284.02 (249.35-322.77) | 574.59 (460.22-708.46) |
| BDHS 2014 | 2010-2013 | 4597 | 38.69 (35.86-41.60) | 24.18 (22.11-26.37) | 62.64 (59.77-65.42) | 255.28 (238.21-282.20) | 523.20 (470.77-580.30) | 733.74 (604.65-887.87) |
| BDHS 2017 | 2014-2016 | 4993 | 49.89 (47.60-52.19) | 33.02 (31.09-35.00) | 66.61 (64.27-68.88) | 357.20 (324.31-394.28) | 751.03 (657.57-856.62) | 843.62 (686.17-1003.0) |

The percentage increase of CS use across several type of health facilities (government, private and non-government) is significantly different (p<0.01, *results not shown*) (Figure 1). In 2004, almost 48% of the CS occurred in government health facilities which was declined to nearly 15% in 2017/18. Reverse trend was reported for CS use in private health facilities, increased from 50% in 2004 to 80% in 2017/18 — a 226% yearly increase between 2004 and 2017/18.

**Figure 1:**
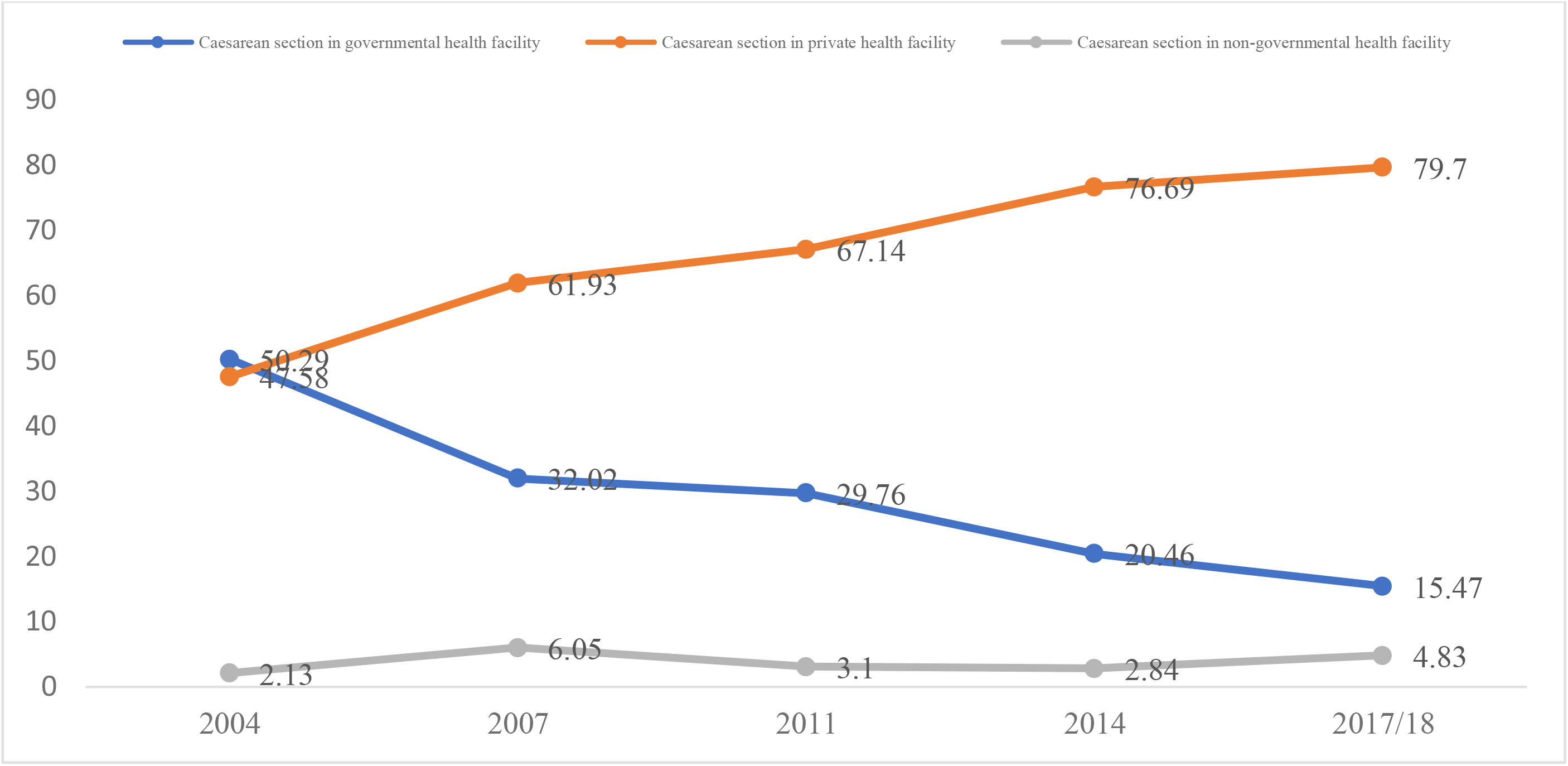
Caesarean section use across type of health facility, Bangladesh 2004-2017/18.

The CS use across major women’s characteristics and place of residence are presented in Figure 2 (a-e). A noticeable change in CS use was reported across wealth quintile, education, working status, and place of residence. In 2004, CS use was mostly prevalent among richest with nearly 70% of the total use where the prevalence of CS use among poorest was <1%. However, with the year passes CS use had been increasing in all wealth quintiles, though, predominantly higher among women with middle to richest wealth quintile with over 78% of the total CS use. The poorest and poorer groups, that represent around one third of the total population with very high fertility rate, utilised only 22% of the total CS use in Bangladesh. Similarly, in all the years, women with secondary and higher education jointly employed over 80% of the total CS whereas remaining 20% CS use was found among illiterate and primary educated women. In 2004, over 61% of the total CS use reported by the women resided in urban area which was declined to 36% in 2017/18. A contradict trend was reported for women resided in rural area, percentage increased from 39% in 2004 to 64% in 2017/18.

**Figure 2(a-d):**
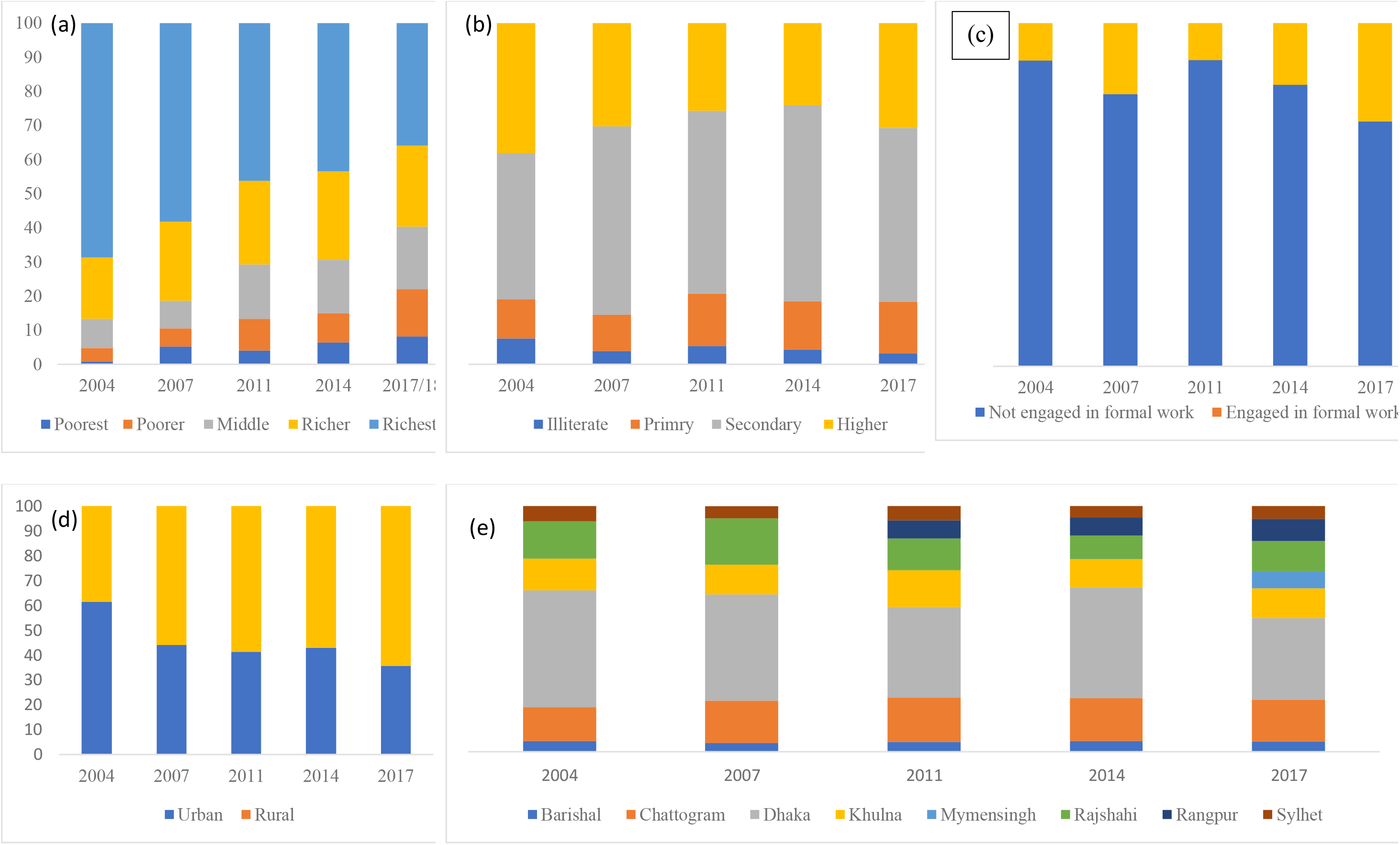
Distribution of Socioeconomic Status among women having caesarean section use in Bangladesh, 2004-2017/18.

Respondents’ CS use were also found to be increased among the disadvantage groups; however, the percentages are found to be varied across the number of disadvantages characteristics that women have had (Figure 3). Around 57% of the women who were in secondary or above education, in the middle to higher wealth quintile and resided in urban area was found to use CS. However, opposite group, that is women who were in lower education class, lower quintile and resided in rural area, were reported to employ only 12% CS use in 2017, which is an increase of only 0.27% in 2004. Around 24% of the women who had at least two disadvantages’ characteristics among three, that is lower education, lower quintile and resided in rural area, reported CS use in 2017, increased from nearly 3% in 2004. A consistent higher use of CS across the survey’s years were found in Dhaka and Khulna divisions.

**Figure 3:**
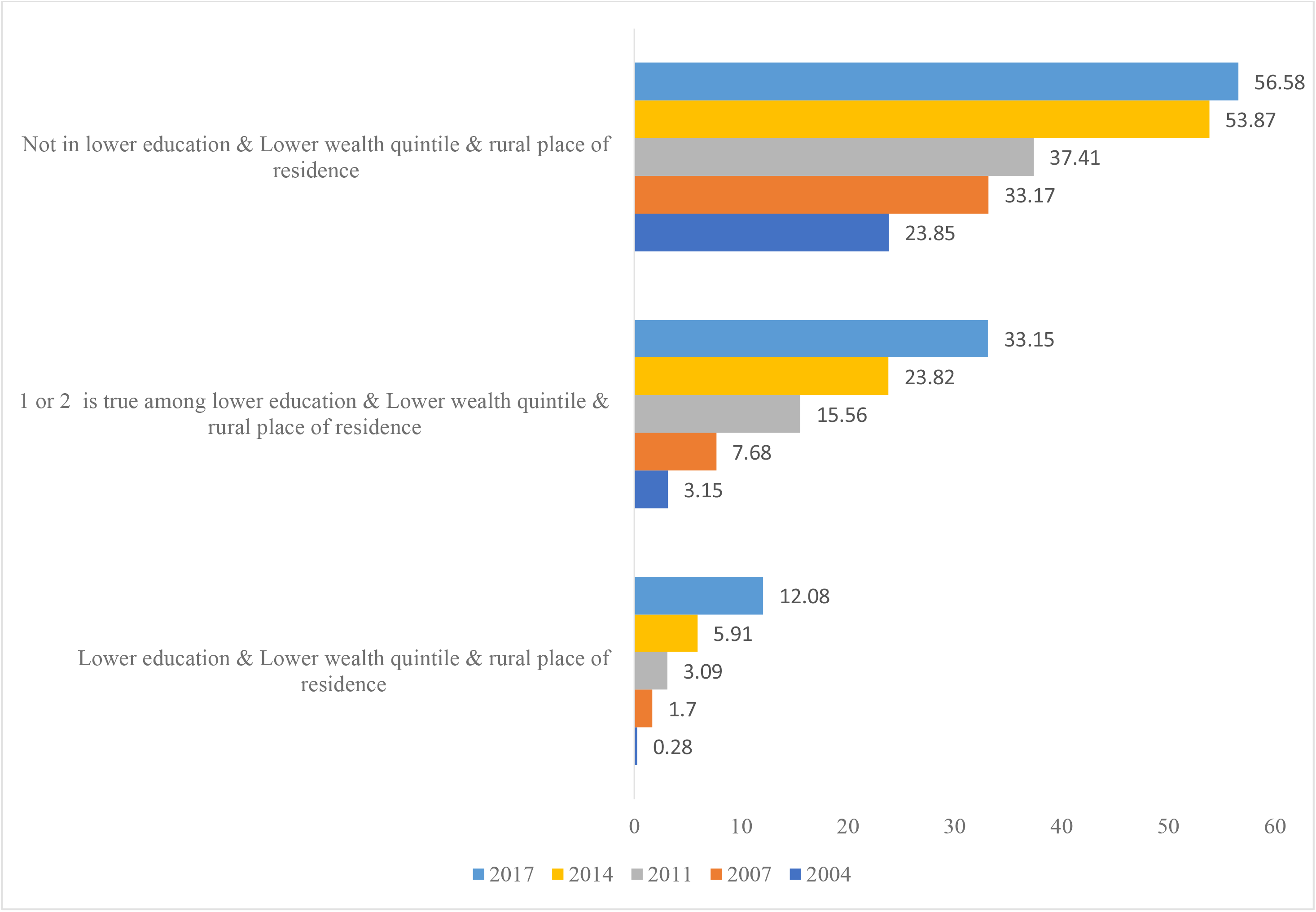
Change in respondents caesarean section used across type of disadvantages characteristics in Bangladesh, 2004-2017/18

The adjusted associations between each of the socio-demographic characteristics and CS use are presented in Table 3. Overall, the risk factors of CS use differed among the survey years and among the different sub-group of socio-demographic characteristics. Increased age, higher education, higher partner education, increase number of antenatal care visits, and improved socio-economic status of the respondent were associated a with higher likelihoods of CS use in all the survey years. Women had more than 2 children and resided in Barishal division were found to be associated with the declined likelihoods of CS use.

**Table 3:**
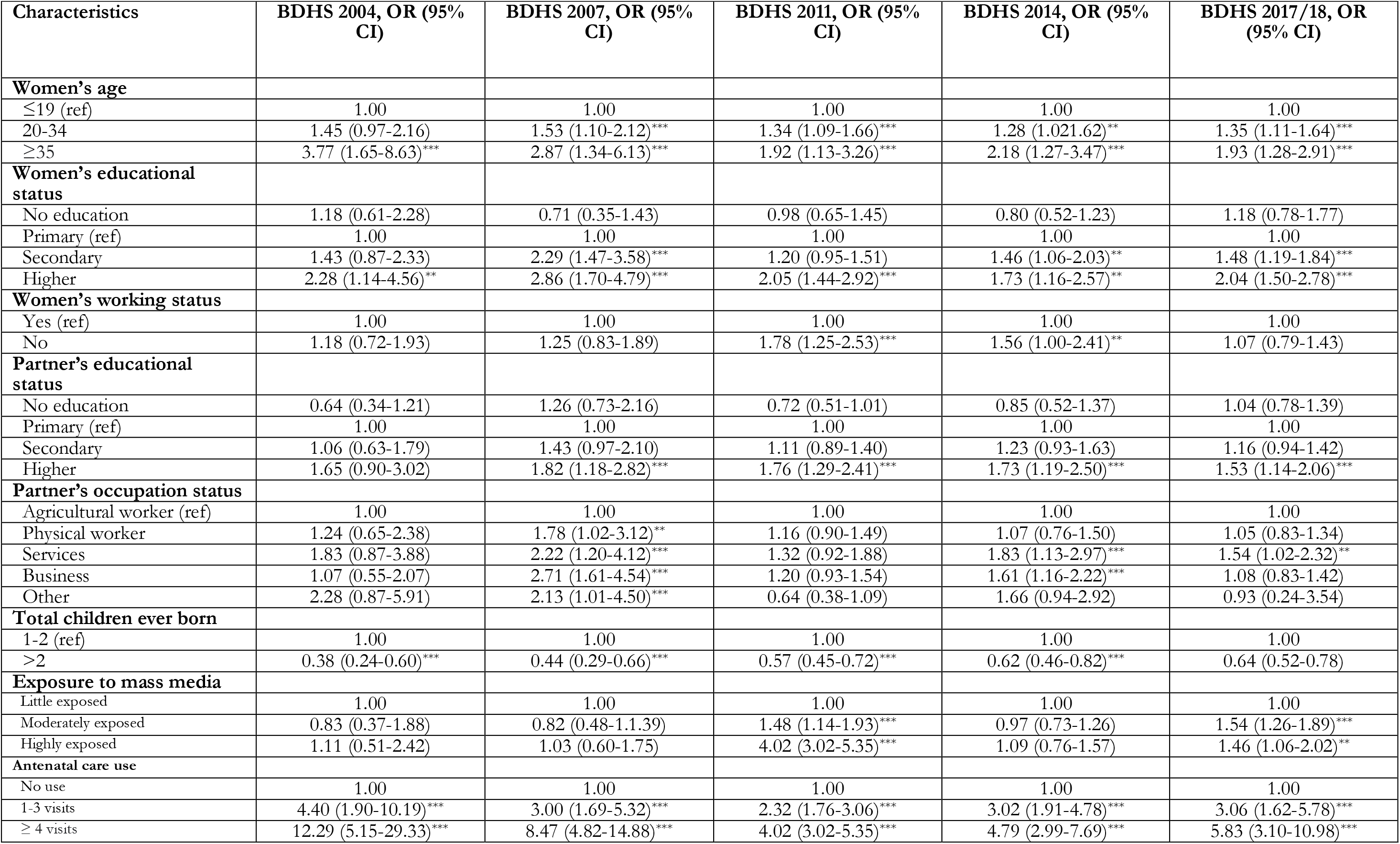
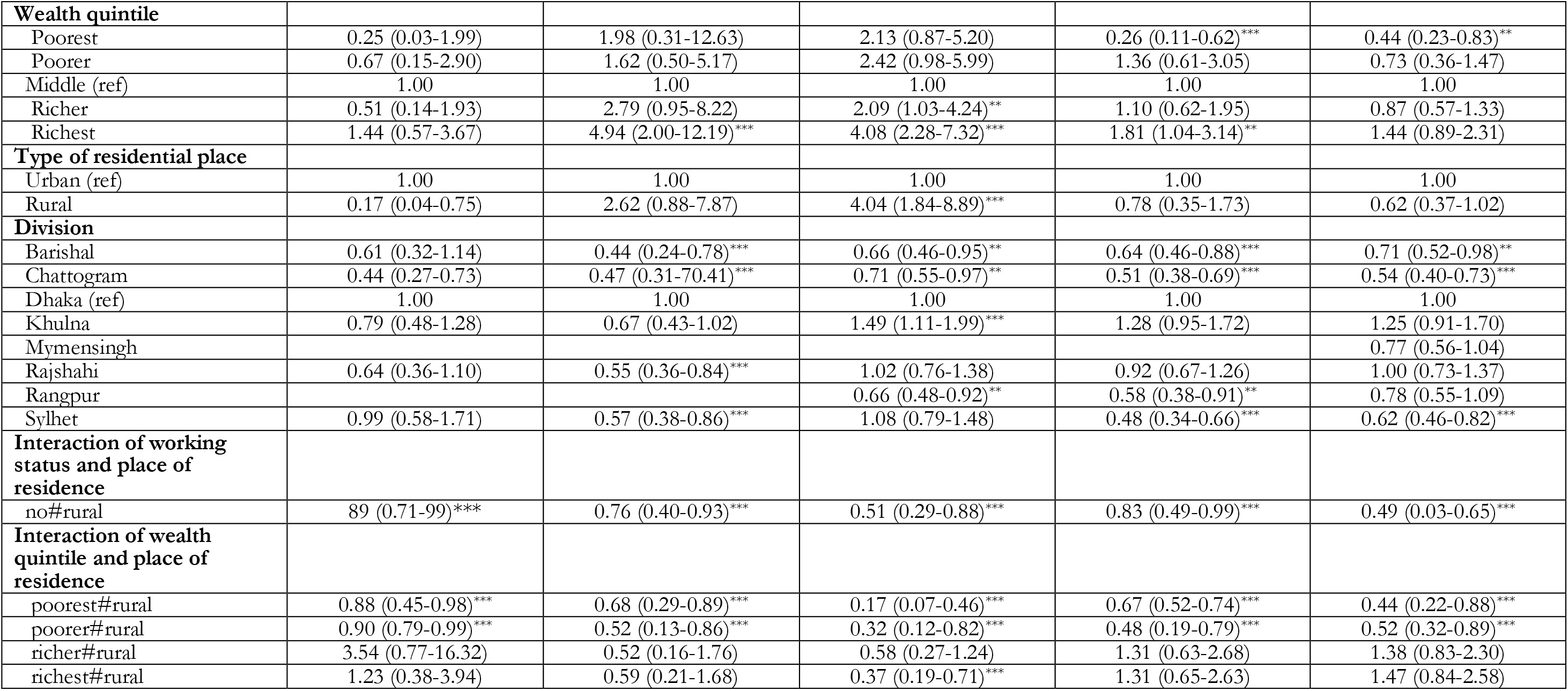
Multilevel logistic regression models for the association of caesarean delivery with socio-demographic characteristics and the interactions of the working status and wealth quintile with place of residence at five-time points (BDHS Surveys: 2004, 2007, 2011, 2014, 2017/18).

| Characteristics | BDHS 2004, OR (95% CI) | BDHS 2007, OR (95% CI) | BDHS 2011, OR (95% CI) | BDHS 2014, OR (95% CI) | BDHS 2017/18, OR (95% CI) |
| --- | --- | --- | --- | --- | --- |
| **Women's age** |  |  |  |  |  |
| ≤19 (ref) | 1.00 | 1.00 | 1.00 | 1.00 | 1.00 |
| 20-34 | 1.45 (0.97-2.16) | 1.53 (1.10-2.12)*** | 1.34 (1.09-1.66)*** | 1.28 (1.021-1.62)** | 1.35 (1.11-1.64)*** |
| ≥35 | 3.77 (1.65-8.63)*** | 2.87 (1.34-6.13)*** | 1.92 (1.13-3.26)*** | 2.18 (1.27-3.47)*** | 1.93 (1.28-2.91)*** |
| **Women's educational status** |  |  |  |  |  |
| No education | 1.18 (0.61-2.28) | 0.71 (0.35-1.43) | 0.98 (0.65-1.45) | 0.80 (0.52-1.23) | 1.18 (0.78-1.77) |
| Primary (ref) | 1.00 | 1.00 | 1.00 | 1.00 | 1.00 |
| Secondary | 1.43 (0.87-2.33) | 2.29 (1.47-3.58)*** | 1.20 (0.95-1.51) | 1.46 (1.06-2.03)** | 1.48 (1.19-1.84)*** |
| Higher | 2.28 (1.14-4.56)** | 2.86 (1.70-4.79)*** | 2.05 (1.44-2.92)** | 1.73 (1.16-2.57)** | 2.04 (1.50-2.78)*** |
| **Women's working status** |  |  |  |  |  |
| Yes (ref) | 1.00 | 1.00 | 1.00 | 1.00 | 1.00 |
| No | 1.18 (0.72-1.93) | 1.25 (0.83-1.89) | 1.78 (1.25-2.53)*** | 1.56 (1.00-2.41)** | 1.07 (0.79-1.43) |
| **Partner's educational status** |  |  |  |  |  |
| No education | 0.64 (0.34-1.21) | 1.26 (0.73-2.16) | 0.72 (0.51-1.01) | 0.85 (0.52-1.37) | 1.04 (0.78-1.39) |
| Primary (ref) | 1.00 | 1.00 | 1.00 | 1.00 | 1.00 |
| Secondary | 1.06 (0.63-1.79) | 1.43 (0.97-2.10) | 1.11 (0.89-1.40) | 1.23 (0.93-1.63) | 1.16 (0.94-1.42) |
| Higher | 1.65 (0.90-3.02) | 1.82 (1.18-2.82)** | 1.76 (1.29-2.41)*** | 1.73 (1.19-2.50)*** | 1.53 (1.14-2.06)*** |
| **Partner's occupation status** |  |  |  |  |  |
| Agricultural worker (ref) | 1.00 | 1.00 | 1.00 | 1.00 | 1.00 |
| Physical worker | 1.24 (0.65-2.38) | 1.78 (1.02-3.12)** | 1.16 (0.90-1.49) | 1.07 (0.76-1.50) | 1.05 (0.83-1.34) |
| Services | 1.83 (0.87-3.88) | 2.22 (1.20-4.12)*** | 1.32 (0.92-1.88) | 1.83 (1.13-2.97)*** | 1.54 (1.02-2.32)** |
| Business | 1.07 (0.55-2.07) | 2.71 (1.61-4.54)*** | 1.20 (0.93-1.54) | 1.61 (1.16-2.22)*** | 1.08 (0.83-1.42) |
| Other | 2.28 (0.87-5.91) | 2.13 (1.01-4.50)*** | 0.64 (0.38-1.09) | 1.66 (0.94-2.92) | 0.93 (0.24-3.54) |
| **Total children ever born** |  |  |  |  |  |
| 1-2 (ref) | 1.00 | 1.00 | 1.00 | 1.00 | 1.00 |
| >2 | 0.38 (0.24-0.60)*** | 0.44 (0.29-0.66)*** | 0.57 (0.45-0.72)*** | 0.62 (0.46-0.82)*** | 0.64 (0.52-0.78) |
| **Exposure to mass media** |  |  |  |  |  |
| Little exposed | 1.00 | 1.00 | 1.00 | 1.00 | 1.00 |
| Moderately exposed | 0.83 (0.37-1.88) | 0.82 (0.48-1.1.39) | 1.48 (1.14-1.93)*** | 0.97 (0.73-1.26) | 1.54 (1.26-1.89)*** |
| Highly exposed | 1.11 (0.51-2.42) | 1.03 (0.60-1.75) | 4.02 (3.02-5.35)*** | 1.09 (0.76-1.57) | 1.46 (1.06-2.02)** |
| **Antenatal care use** |  |  |  |  |  |
| No use | 1.00 | 1.00 | 1.00 | 1.00 | 1.00 |
| 1-3 visits | 4.40 (1.90-10.19)*** | 3.00 (1.69-5.32)*** | 2.32 (1.76-3.06)*** | 3.02 (1.91-4.78)*** | 3.06 (1.62-5.78)*** |
| ≥ 4 visits | 12.29 (5.15-29.33)*** | 8.47 (4.82-14.88)*** | 4.02 (3.02-5.35)*** | 4.79 (2.99-7.69)*** | 5.83 (3.10-10.98)*** |
| **Wealth quintile** |  |  |  |  |  |
| Poorest | 0.25 (0.03-1.99) | 1.98 (0.31-12.63) | 2.13 (0.87-5.20) | 0.26 (0.11-0.62)*** | 0.44 (0.23-0.83)** |
| Poorer | 0.67 (0.15-2.90) | 1.62 (0.50-5.17) | 2.42 (0.98-5.99) | 1.36 (0.61-3.05) | 0.73 (0.36-1.47) |
| Middle (ref) | 1.00 | 1.00 | 1.00 | 1.00 | 1.00 |
| Richer | 0.51 (0.14-1.93) | 2.79 (0.95-8.22) | 2.09 (1.03-4.24)** | 1.10 (0.62-1.95) | 0.87 (0.57-1.33) |
| Richest | 1.44 (0.57-3.67) | 4.94 (2.00-12.19)*** | 4.08 (2.28-7.32)*** | 1.81 (1.04-3.14)** | 1.44 (0.89-2.31) |
| **Type of residential place** |  |  |  |  |  |
| Urban (ref) | 1.00 | 1.00 | 1.00 | 1.00 | 1.00 |
| Rural | 0.17 (0.04-0.75) | 2.62 (0.88-7.87) | 4.04 (1.84-8.89)*** | 0.78 (0.35-1.73) | 0.62 (0.37-1.02) |
| **Division** |  |  |  |  |  |
| Barishal | 0.61 (0.32-1.14) | 0.44 (0.24-0.78)*** | 0.66 (0.46-0.95)** | 0.64 (0.46-0.88)*** | 0.71 (0.52-0.98)** |
| Chattogram | 0.44 (0.27-0.73) | 0.47 (0.31-0.70)*** | 0.71 (0.55-0.97)** | 0.51 (0.38-0.69)*** | 0.54 (0.40-0.73)*** |
| Dhaka (ref) | 1.00 | 1.00 | 1.00 | 1.00 | 1.00 |
| Khulna | 0.79 (0.48-1.28) | 0.67 (0.43-1.02) | 1.49 (1.11-1.99)*** | 1.28 (0.95-1.72) | 1.25 (0.91-1.70) |
| Mymensingh |  |  |  |  | 0.77 (0.56-1.04) |
| Rajshahi | 0.64 (0.36-1.10) | 0.55 (0.36-0.84)*** | 1.02 (0.76-1.38) | 0.92 (0.67-1.26) | 1.00 (0.73-1.37) |
| Rangpur |  |  | 0.66 (0.48-0.92)** | 0.58 (0.38-0.91)** | 0.78 (0.55-1.09) |
| Sylhet | 0.99 (0.58-1.71) | 0.57 (0.38-0.86)*** | 1.08 (0.79-1.48) | 0.48 (0.34-0.66)*** | 0.62 (0.46-0.82)*** |
| **Interaction of working status and place of residence** |  |  |  |  |  |
| no#rural | 89 (0.71-99)*** | 0.76 (0.40-0.93)*** | 0.51 (0.29-0.88)*** | 0.83 (0.49-0.99)*** | 0.49 (0.03-0.65)*** |
| **Interaction of wealth quintile and place of residence** |  |  |  |  |  |
| poorest#rural | 0.88 (0.45-0.98)*** | 0.68 (0.29-0.89)*** | 0.17 (0.07-0.46)*** | 0.67 (0.52-0.74)*** | 0.44 (0.22-0.88)*** |
| poorer#rural | 0.90 (0.79-0.99)*** | 0.52 (0.13-0.86)*** | 0.32 (0.12-0.82)*** | 0.48 (0.19-0.79)*** | 0.52 (0.32-0.89)*** |
| richer#rural | 3.54 (0.77-16.32) | 0.52 (0.16-1.76) | 0.58 (0.27-1.24) | 1.31 (0.63-2.68) | 1.38 (0.83-2.30) |
| richest#rural | 1.23 (0.38-3.94) | 0.59 (0.21-1.68) | 0.37 (0.19-0.71)*** | 1.31 (0.65-2.63) | 1.47 (0.84-2.58) |

From the interaction terms, women’s rural place of residence and their engagement in formal job were associated with reduced likelihoods of CS use in all the surveys. In 2004, this interaction was found to be associated with 11% (OR, 0.89, 95% CI, 0.71-0.99) reduction of CS use which was found to be increased to 51% (OR, 0.49, 0.03-0.65) reduction in 2017/18. Similarly, consistent lower likelihoods of CS use were found among rural poorest and poorer women with a rising trend of declining over the years. In 2004, 12% (OR, 0.45-0.98) declined likelihoods of CS use was found among rural poorest women. This likelihood was 66% (OR,44, 95% CI, 0.22-0.88) lower in the 2017/18 survey. Comparably, around 10% (OR, 0.90, 95% CI, 0.79-0.99) lower likelihood of CS use was found among rural poorer women in 2004 survey, which increased to 48% (OR, 0.52, 95% CI, 0.32-0.89) declined in the 2017/18 survey.

## Discussion

Nearly one-third of the total women in Bangladesh use CS as per the 2017/18 survey which is around 751% higher than the 2004 survey of 3.88% CS use. Private health facilities had been popular in providing the CS delivery over the survey years and now providing nearly 80% of the total CS in Bangladesh. Alternatively, the share of the government health facilities in providing CS declined substantially, from 50.29% in 2004 to 15.47% in 2017/18. Private health facilities in Bangladesh are mostly profit ambitious and located in urban centre. Consequently, respondents resided in rural area, that cover over 70% of the total population in Bangladesh, with poor economic condition could not access this service — as this study reports. This indicates a group of women in Bangladesh use CS abusively while other group could not access this service even under necessary condition. As a result, they are more likely to face short-term and long-term health consequences, problem in the subsequent pregnancies, as well as death from obstetric complications. Efforts are needed to prevent medically unnecessary CS use along with guaranteed access to save delivery for rural and poor women.

This study reported a rapid rise on CS use in Bangladesh in general and in private health facilities in particular. The use of CS declined substantially in the government health facilities. This trend contradicts to what have been reported worldwide and LMICs [27], including India [28], Pakistan [29] and Ethiopia [30]. Though such exponential rising rate of CS in private health facilities in Bangladesh has two different roots, both from the individual and health facilities level, however, the government poor regulation on the private health facilities is the major factor.

Over the decade, CS use has been rising exponentially and becoming popular in Bangladesh, mainly because of its increasing availability in the health facilities on women’s demand without medical necessity [31]. Women are also misguided by the healthcare personnel and their peers who had undergone through CS with wrong insight over this method [10]. They often consider CS use as a way of reducing labor pain and safe approach for both women and upcoming newborn, even where there were no medical necessity [21, 32]. This is the primary reason of booming the CS use in the private health facilities as CS use on women’s demand is mostly available there – contradict to the government health facilities practice [18]. Moreover, a large percentage of women admitted in the government health centre, finally changes their mind and shift to the private facilities and choose to undergone through the CS [33, 34]. The reasons are lack of healthcare personnel in the governmental health facilities, poor quality of care, long waiting time for the doctors and unsupportive behaviours of the nurses and other associated medical staffs [18, 34]. Conversely, in the private health facilities, even though they are profit making institutions, they always ensure better care and concern for the patients[18]. The brokers are also active in the governmental health facilities independently and/or in cooperation with the corrupt healthcare personnel to motivate women to shift to the private health facilities by promising the advantages and better care provided by the CS [35]. About 15% use of CS in the government health facilities are mainly because of their main role on handling more complicated cases who are directly admitted there as well as referred from the private and non-governmental health facilities [18].

It is found in this study that such rising rate of CS as well as increase familiarization of private facilities for CS contribute to occurring double burden of CS in Bangladesh. A similar trend is found in other LMICs; however, the strength is much lower than Bangladesh [36-38]. The average cost of performing CS in the private health facilities in Bangladesh is 612 USD, twice higher than the average monthly income (USD 301) of Bangladeshi population and 6 to 8 fold higher than the Bangladeshi poorer population [5, 39]. For rural women, such higher cost is accompanied with transportation and relocation costs to urban area where all private health facilities and majority of the government health facilities are located. Therefore, poorer women in general and rural poor women in particular could not use this service. These increase the rate of home delivery, particularly among the rural women who usually have higher number of children [9, 40]. In this case, most women depend on their experienced from the previous pregnancies rather than accessing services including antenatal care, which is the motivator of delivery care access. Our findings are supportive to this conclusion and it is similar to what has been reported in other LMICs [29, 33, 36, 38]. Even if the use of CS is required, the rural women, particularly those who have no proper income and belong to the poor socio-economic condition, often need to sell their properties, or borrow money from the usurer [31]. The findings of the interaction effects of place of residence with women of no formal job engagement and poor wealth quintile have been justified through these linkages. Such burden is even intensified because of the government single-mindedness to reduce the number of CS rather than considering the justification of its importance and requirements, particularly among the disadvantage groups. However, this issue may not be true for the urban poor women as the non-governmental health facilities there provide free services or with a very minimum cost [11].

The governmental policy and programs to control the CS use are also not properly effective. Instead of decreasing, it is often lead to the increase or double the burden of CS use. In 2019 a committee of experts and stakeholders was set up following the High Court order to stop unnecessary CS use [31]. The committee later introduced a system whereby all health facilities were to report history of all deliveries carried out by filling a form, either normal or CS. For CS, the reasons of choosing and conducting CS were also need to be reported. These procedures contradict to what is recommended by the International Federation of Gynaecology and Obstetrics for control CS: imposing the fixed fee for live birth, obliging hospital to publish their statistics and fully informing women about the risks of CS use [41]. The government also does not impose any restriction on private facilities to provide delivery care along with CS as part of corporate social responsibility or based on mutual agreement with the government where the government will support for CS for the disadvantages’ groups. No programs are also carried out to provide delivery services along with CS requirement at the rural areas. For instance, the government does not take any policies and programs to ensure CS at the community clinics or union health complex, though they are higher in number, located in rural areas for every 6000 population and currently playing a major role to provide antenatal care and postnatal care [42]. Consequently, the current programs to control CS does not work properly in either way, controlling CS as well as reducing the double burden of CS. These suggest urgent need for the policies and programs from the top country level to effectively regulate the private health facilities on CS use as well as ensuring accessing to CS for those who do not afford or have the proper financial support.

This study has a number of strengths and limitations. The major limitation is the analysis of cross-sectional data; therefore, all findings are correlational and not casual. Moreover, the data were collected retrospectively which indicate a possibility of recall error, particularly for the variable like antenatal care use. In addition, since the data on the CS medical requirement is not available, our explanations about the double burden are based on probability rather than considering whether such use is medically justified. However, these interaction effects will enable policy makers to identify the group of women who could not access this service and develop policies and programs accordingly. Furthermore, other than the factors adjusted in the model, there are many other factors including transportation and quality of the care which are also important determinant of CS use. We could not adjust them in the model because of lack of data. Regardless of these limitations, this study is the first of its kind in Bangladesh and other LMICs that bring the issue of double burden of CS into focus. The factors associated with the CS along with the interaction effects of the women’s disadvantage characteristics are also determined. This will enable the policy makers to plan for more effective policies and programs.

## Conclusions

This study reported a rapid rise on CS over the survey years, from nearly 4% in 2004 to 33% in 2017/18. Nearly 80% of the CS are performed in the private health facilities, increased from about 48% in 2004. In contrary, CS use declined from nearly 50% in 2004 to only 15% in 2017/18 in the governmental health facilities. Such rising familiarization of CS in the private health facilities could lead to double burden of CS in Bangladesh. Since private health facilities are profit ambitious, women with improved socio-economic condition are the main clients to this service. On the other hand, rural, not working, and poorer women cannot afford these services. This goes along with the government health facilities level challenges including lack of healthcare personnel and poor quality of care, restrict disadvantages women in accessing CS even when it is for medical requirement. This indicates a pathway increasing short term and long-term obstetric and medical complications as well as deaths from obstetric complications. Therefore, government regulations are strictly required in order to control CS use in general and private health facilities in particular. The government should also impose regulations on the private health facilities to deliver CS when required medically among disadvantages with minimum fees or even free of charge when necessary.

## Data Availability

The data sets used and analysed in this study are available from the Measure DHS website: https://dhsprogram.com/data/available-datasets.cfm

## Acknowledgement

Special thanks to the MEASURE DHS from the authors for granting access to the 2017/18 BDHS data.

## Authors’ contributions

MNK designed the study, performed the data analysis, and wrote the first draft of this manuscript. MMR, Kabir MA, and AAS critically reviewed and edited the previous versions of this manuscript. All authors approved this final version of the manuscript.

---

STROBE Statement—Checklist of items that should be included in reports of ***cross-sectional studies***

|  | Item No | Recommendation | Page No |
| --- | --- | --- | --- |
| Title and abstract | 1 | (a) Indicate the study’s design with a commonly used term in the title or the abstract | 2 |
|  |  | (b) Provide in the abstract an informative and balanced summary of what was done and what was found | 2-3 |
| Introduction |  |  |  |
| Background/rationale | 2 | Explain the scientific background and rationale for the investigation being reported | 4/5 |
| Objectives | 3 | State specific objectives, including any prespecified hypotheses | 5 |
| Methods |  |  |  |
| Study design | 4 | Present key elements of study design early in the paper | 5 |
| Setting | 5 | Describe the setting, locations, and relevant dates, including periods of recruitment, exposure, follow-up, and data collection | 6 |
| Participants | 6 | (a) Give the eligibility criteria, and the sources and methods of selection of participants | 6/7 |
| Variables | 7 | Clearly define all outcomes, exposures, predictors, potential confounders, and effect modifiers. Give diagnostic criteria, if applicable | 6/7 |
| Data sources/measurement | 8* | For each variable of interest, give sources of data and details of methods of assessment (measurement). Describe comparability of assessment methods if there is more than one group | 6/7 |
| Bias | 9 | Describe any efforts to address potential sources of bias |  |
| Study size | 10 | Explain how the study size was arrived at | 7 |
| Quantitative variables | 11 | Explain how quantitative variables were handled in the analyses. If applicable, describe which groupings were chosen and why | 7 |
| Statistical methods | 12 | (a) Describe all statistical methods, including those used to control for confounding | 7 |
|  |  | (b) Describe any methods used to examine subgroups and interactions | 7 |
|  |  | (c) Explain how missing data were addressed | 7 |
|  |  | (d) If applicable, describe analytical methods taking account of sampling strategy | 7 |
|  |  | (e) Describe any sensitivity analyses | 7 |
| Results |  |  |  |
| Participants | 13* | (a) Report numbers of individuals at each stage of study—eg numbers potentially eligible, examined for eligibility, confirmed eligible, included in the study, completing follow-up, and analysed | 7/9 |
|  |  | (b) Give reasons for non-participation at each stage | 7/9 |
|  |  | (c) Consider use of a flow diagram | 7/9 |
| Descriptive data | 14* | (a) Give characteristics of study participants (eg demographic, clinical, social) and information on exposures and potential confounders | 7/9 |
|  |  | (b) Indicate number of participants with missing data for each variable of interest | 7/9 |
| Outcome data | 15* | Report numbers of outcome events or summary measures | 7/9 |
| Main results | 16 | (a) Give unadjusted estimates and, if applicable, confounder-adjusted estimates and their precision (eg, 95% confidence interval). Make clear which confounders were adjusted for and why they were included | 7/9 |
|  |  | (b) Report category boundaries when continuous variables were categorized | 7/9 |
|  |  | (c) If relevant, consider translating estimates of relative risk into absolute risk for a meaningful time period | 7/9 |
| Other analyses | 17 | Report other analyses done—eg analyses of subgroups and interactions, and sensitivity analyses | 7/9 |
| **Discussion** |  |  |  |
| Key results | 18 | Summarise key results with reference to study objectives | 10 |
| Limitations | 19 | Discuss limitations of the study, taking into account sources of potential bias or imprecision. Discuss both direction and magnitude of any potential bias | 13 |
| Interpretation | 20 | Give a cautious overall interpretation of results considering objectives, limitations, multiplicity of analyses, results from similar studies, and other relevant evidence | 10/13 |
| Generalisability | 21 | Discuss the generalisability (external validity) of the study results | 10/13 |
| **Other information** |  |  |  |
| Funding | 22 | Give the source of funding and the role of the funders for the present study and, if applicable, for the original study on which the present article is based | 14 |

\*Give information separately for exposed and unexposed groups.
**Note:** An Explanation and Elaboration article discusses each checklist item and gives methodological background and published examples of transparent reporting. The STROBE checklist is best used in conjunction with this article (freely available on the Web sites of PLoS Medicine at http://www.plosmedicine.org/, Annals of Internal Medicine at http://www.annals.org/, and Epidemiology at http://www.epidem.com/). Information on the STROBE Initiative is available at www.strobe-statement.org.

## Reference

1. Caesarean section rates continue to rise, amid growing inequalities in access: WHO [Internet]. World Health Organization,. 2021

2. Dumont A, Guilmoto CZ. Too many yet too few: The double burden of Caesarean births. Population Societies. 2020;(9):1–4.

3. Betran AP, Ye J, Moller A-B, Souza JP, Zhang J. Trends and projections of caesarean section rates: global and regional estimates. BMJ Global Health. 2021;6(6):e005671.

4. Betran A, Torloni MR, Zhang J, Gülmezoglu A, Section WWGoC, Aleem H, et al. WHO statement on caesarean section rates. BJOG: An International Journal of Obstetrics & Gynaecology. 2016;123(5):667–70.

5. Haider MR, Rahman MM, Moinuddin M, Rahman AE, Ahmed S, Khan MM. Ever-increasing Caesarean section and its economic burden in Bangladesh. PloS one. 2018;13(12):e0208623.

6. Sway A, Nthumba P, Solomkin J, Tarchini G, Gibbs R, Ren Y, et al. Burden of surgical site infection following cesarean section in sub-Saharan Africa: a narrative review. International journal of women’s health. 2019;11:309.

7. Beogo I, Rojas BM, Gagnon M-P. Determinants and materno-fetal outcomes related to cesarean section delivery in private and public hospitals in low-and middle-income countries: a systematic review and meta-analysis protocol. Systematic reviews. 2017;6(1):1–6.

8. Keag OE, Norman JE, Stock SJ. Long-term risks and benefits associated with cesarean delivery for mother, baby, and subsequent pregnancies: Systematic review and meta-analysis. PLoS medicine. 2018;15(1):e1002494.

9. Khan MN, Islam MM, Shariff AA, Alam MM, Rahman MM. Socio-demographic predictors and average annual rates of caesarean section in Bangladesh between 2004 and 2014. PloS one. 2017;12(5):e0177579.

10. Khan MN, Islam MM, Rahman M. Inequality in utilization of cesarean delivery in Bangladesh: a decomposition analysis using nationally representative data. Public health. 2018;157:111–20.

11. Khan MN AS, Rahman MM. Influence of health facility level factors on raising caesarean section delivery in Bangladesh: Evidence from linked data of population and health facility survey. MedRxiv. 2021.

12. Tegegne TK, Chojenta C, Getachew T, Smith R, Loxton D. Caesarean Delivery Use in Ethiopia: A Spatial and Hierarchical Bayesian Analysis. 2021.

13. Antoine C, Young BK. Cesarean section one hundred years 1920–2020: the Good, the Bad and the Ugly. Journal of Perinatal Medicine. 2021;49(1):5–16.

14. Chowdhury HA, Ahmed KR, Jebunessa F, Akter J, Hossain S, Shahjahan M. Factors associated with maternal anaemia among pregnant women in Dhaka city. BMC women’s health. 2015;15(1):1–6.

15. Hyder SZ, Persson L-Å, Chowdhury M, Lönnerdal B, Ekström E-C. Anaemia and iron deficiency during pregnancy in rural Bangladesh. Public health nutrition. 2004;7(8):1065–70.

16. Rahman MA, Rahman MS, Aziz Rahman M, Szymlek-Gay EA, Uddin R, Islam SMS. Prevalence of and factors associated with anaemia in women of reproductive age in Bangladesh, Maldives and Nepal: Evidence from nationally-representative survey data. Plos one. 2021;16(1):e0245335.

17. Boatin AA, Schlotheuber A, Betran AP, Moller A-B, Barros AJ, Boerma T, et al. Within country inequalities in caesarean section rates: observational study of 72 low and middle income countries. bmj. 2018;360.

18. Mia MN, Islam MZ, Chowdhury MR, Razzaque A, Chin B, Rahman MS. Socio-demographic, health and institutional determinants of caesarean section among the poorest segment of the urban population: Evidence from selected slums in Dhaka, Bangladesh. SSM-population health. 2019;8:100415.

19. Neuman M, Alcock G, Azad K, Kuddus A, Osrin D, More NS, et al. Prevalence and determinants of caesarean section in private and public health facilities in underserved South Asian communities: cross-sectional analysis of data from Bangladesh, India and Nepal. BMJ open. 2014;4(12):e005982.

20. Assembly G. Sustainable development goals. SDGs Transform Our World. 2015;2030.

21. National Institute of Population Research and Training (NIPORT) and ICF. Bangladesh Demographic and Health Survey 2017. Dhaka, Bangladesh: NIPORT, ACPR, and ICF.: 2021.

22. National Institute of Population Research and Training (NIPORT) and ICF. Bangladesh Demographic and Health Survey 2014. Dhaka, Bangladesh: NIPORT, ACPR, and ICF.: 2017.

23. National Institute of Population Research and Training (NIPORT) and ICF. Bangladesh Demographic and Health Survey 2004. Dhaka, Bangladesh: NIPORT, ACPR, and ICF.: 2005.

24. National Institute of Population Research and Training (NIPORT) and ICF. Bangladesh Demographic and Health Survey 2011. Dhaka, Bangladesh: NIPORT, ACPR, and ICF.: 2012.

25. National Institute of Population Research and Training (NIPORT) and ICF. Bangladesh Demographic and Health Survey 2007. Dhaka, Bangladesh: NIPORT, ACPR, and ICF.: 2007.

26. Modugno L, Monari P, Giannerini S, Cagnone S. A multilevel approach for repeated cross-sectional data.

27. Betrán AP, Ye J, Moller A-B, Zhang J, Gülmezoglu AM, Torloni MR. The increasing trend in caesarean section rates: global, regional and national estimates: 1990-2014. PloS one. 2016;11(2):e0148343.

28. Sengupta A, Sagayam MS, Reja T. Increasing trend of C-section deliveries in India: A comparative analysis between southern states and rest of India. Sexual & Reproductive Healthcare. 2021;28:100608.

29. Mumtaz S, Bahk J, Khang Y-H. Rising trends and inequalities in cesarean section rates in Pakistan: Evidence from Pakistan Demographic and Health Surveys, 1990-2013. PloS one. 2017;12(10):e0186563.

30. Shibre G, Idriss-Wheeler D, Bishwajit G, Yaya S. Observed trends in the magnitude of socioeconomic and area-based inequalities in use of caesarean section in Ethiopia: a cross-sectional study. BMC Public Health. 2020;20(1):1–12.

31. Bangladesh: 51 percent increase in “unnecessary” C-section in two years [Internet]. 2018.

32. National Institute of Population Research and Training (NIPORT) and ICF. Bangladesh Health Facility Survey 2017. Dhaka, Bangladesh: NIPORT, ACPR, and ICF.: 2019.

33. Vieira GO, Fernandes LG, de Oliveira NF, Silva LR, de Oliveira Vieira T. Factors associated with cesarean delivery in public and private hospitals in a city of northeastern Brazil: a cross-sectional study. BMC pregnancy and childbirth. 2015;15(1):1–9.

34. Sk R. Does delivery in private hospitals contribute largely to Caesarean Section births? A path analysis using generalised structural equation modelling. PloS one. 2020;15(10):e0239649.

35. Mridha MK, Anwar I, Koblinsky M. Public-sector maternal health programmes and services for rural Bangladesh. Journal of health, population, and nutrition. 2009;27(2):124.

36. Singh P, Hashmi G, Swain PK. High prevalence of cesarean section births in private sector health facilities-analysis of district level household survey-4 (DLHS-4) of India. BMC public health. 2018;18(1):1–10.

37. Bhatia M, Banerjee K, Dixit P, Dwivedi LK. Assessment of variation in cesarean delivery rates between public and private health facilities in India from 2005 to 2016. JAMA network open. 2020;3(8):e2015022–e.

38. Melesse MB, Geremew AB, Abebe SM. High prevalence of caesarean birth among mothers delivered at health facilities in Bahir Dar city, Amhara region, Ethiopia. A comparative study. PloS one. 2020;15(4):e0231631.

39. Bangladesh Bureau of Statistics (BBS). Bangladesh Statistics 2019. Dhaka, Bangladesh. : Bangladesh Bureau of Statistics (BBS) Statistics and Informatics Division (SID) Ministry of Planning, 2019.

40. Khan MN, Harris ML, Loxton D. Does unintended pregnancy have an impact on skilled delivery care use in Bangladesh? A nationally representative cross-sectional study using Demography and Health Survey data. Journal of Biosocial Science. 2020:1–17.

41. Sheiner E, Kapur A, Retnakaran R, Hadar E, Poon LC, McIntyre HD, et al. FIGO (International Federation of Gynecology and Obstetrics) Postpregnancy Initiative: Long-term Maternal Implications of Pregnancy Complications—Follow-up Considerations. International Journal of Gynecology & Obstetrics. 2019;147:1–31.

42. Khan MN, Harris ML, Loxton D. Assessing the effect of pregnancy intention at conception on the continuum of care in maternal healthcare services use in Bangladesh: Evidence from a nationally representative cross-sectional survey. PloS one. 2020;15(11):e0242729.

